# Hypothalamic subunit volumes in schizophrenia and bipolar spectrum disorders

**DOI:** 10.1101/2023.05.31.23289245

**Authors:** Aurora Ruggeri, Stener Nerland, Lynn Mørch-Johnsen, Kjetil Nordbø Jørgensen, Claudia Barth, Laura Wortinger, Dimitrios Andreou, Ole A. Andreassen, Ingrid Agartz

## Abstract

**Background:** The hypothalamus is central to many hormonal and autonomous nervous system pathways. Emerging evidence indicates that these pathways may be disrupted in schizophrenia and bipolar disorder. Yet, few studies have examined the volumes of hypothalamic subunits in these patient groups. We compared hypothalamic subunit volumes in individuals with psychotic disorders to healthy controls.

**Study Design:** We included 344 patients with schizophrenia spectrum disorders (SCZ), 340 patients with bipolar disorders (BPD) and 684 age-and-sex-matched healthy controls (CTR). Total hypothalamus and five hypothalamic subunit volumes were extracted from T1-weighted MRI using an automated Bayesian segmentation method. Regression models, corrected for age, age^2^, sex, and segmentation-based intracranial volume (sbTIV), were used to examine diagnostic group differences, interactions with sex, and associations with clinical symptoms, antipsychotic medication, antidepressants and mood stabilizers.

**Study Results:** SCZ had larger volumes in the left inferior tubular subunit and smaller right anterior inferior, right anterior superior and right posterior hypothalamic subunits compared to CTR. BPD did not differ significantly from CTR for any hypothalamic subunit volume, however, there was a significant sex-by-diagnosis interaction. Analyses stratified by sex showed smaller right hypothalamus and right posterior subunit volumes in male patients, but not female patients, relative to same-sex controls. There was a significant association between BPD currently taking antipsychotic medication and the left inferior tubular subunits volumes.

**Conclusions:** Our results show regional-specific alterations in hypothalamus subunit volumes in individuals with SCZ, with relevance to HPA-axis dysregulation, circadian rhythm disruption and cognition impairment.

**Highlights:**

- Smaller right anterior-inferior, right anterior superior, right posterior and larger left inferior tubular subunit volumes were observed in individuals with schizophrenia spectrum disorders compared to healthy controls.
- Smaller total right hypothalamus volume and smaller right posterior subunit volumes were found for males, but not females, with bipolar disorders.
- There were no significant associations with clinical symptom dimensions.
- Larger left inferior tubular subunits volumes were observed in individuals with bipolar disorders currently taking antipsychotic medication.

## Introduction

The hypothalamus, a small brain structure located above the midbrain and below the thalamus, is comprised of distinct nuclei that are vital for maintaining body homeostasis and physiological functions such as body temperature, the sleep-wake cycle, thirst, and hunger. Together with the pituitary gland, the hypothalamus acts as a key regulator of the neuroendocrine system. It plays an important role in the hypothalamus-pituitary-adrenal (HPA) axis, which is thought to be affected in mood and psychotic disorders^1,2,3,4,5^.

Morphological deficits in the hypothalamus have been reported in severe mental disorders, including in schizophrenia spectrum disorder (SCZ) and bipolar spectrum disorder (BPD). However, previous neuroimaging studies of total hypothalamus volumes in SCZ and BPD have yielded conflicting results, where most studies have relied on small samples. In SCZ studies both larger^6,7,8^ and smaller^9^ volumes compared to healthy controls have been reported, whereas other studies have reported no differences^10^. In BPD, larger total hypothalamus volume in patients compared to healthy controls has been reported^5^; however, this contrasts with findings of smaller volumes from post-mortem studies^11^.

Within the hypothalamus, individual nuclei exert different endocrine and autonomic functions. The main nuclei are the suprachiasmatic nucleus (SCN), ventromedial nucleus (VMN); paraventricular nucleus (PVN), preoptic nucleus, supraoptic nucleus (SON), arcuate nucleus (ARC), and the mammillary bodies. The SCN^12,13^ regulates the circadian rhythm and sleep; both processes which are disrupted in SCZ and BPD disorders^14^. The VMN is important for the regulation of female sexual behavior, satiety and caloric intake, energy balance and cardiovascular rhythm^15,16^. Metabolic abnormalities have been observed among individuals with SCZ^17^. Production of the peptides oxytocin and vasopressin occurs in the paraventricular and supraoptic nuclei (SON)^18,19^. Oxytocin plays a critical role in social cognition and several studies have examined the potential clinical benefits of oxytocin for improvement of psychopathology in SCZ^20,21^. The preoptic nucleus is involved in the regulation of sleep, body temperature and modulates food intake in response to temperature change^22,23^. The ARC is an important site for the integration of peripheral signals with other sensory information and for the syntheses of dopamine and growth-releasing hormone, as well as for relaying leptin signals to other parts of the hypothalamus^24^. Dopamine is a modulatory neurotransmitter involved in the pathology of SCZ ^25,26^ and in the control of prolactin release from the anterior pituitary^27^. Disrupted homoeostasis between the dopamine transporter and dopamine receptors has also been reported in depressive phases of BPD^28^. Mammillary bodies are part of the Papez Circuit, and are involved in memory recollection, including spatial and episodic memory consolidation^29,30^. Impaired memory recollection constitutes a potential cognitive endophenotype in SCZ^31^ and in subsyndromal individuals with BPD^32^.

Conflicting results have been reported in previous magnetic resonance imaging (MRI) and postmortem studies on individual hypothalamic nuclei volumes. In SCZ, both larger^6,7^ and similar size^33^ mammillary body volumes have been observed *in vivo* compared to healthy controls. In post-mortem studies of individuals with SCZ significant differences of the SON, PVN or the SCN have not been observed, while a larger PVN volume was reported in an *in vivo* MRI study^6^.

Whether or not variation in hypothalamic nuclei volumes depends on patient characteristics is largely unknown. A sex-effect on the volumes of the mammillary bodies has been reported^10^, with larger volumes in male compared to female individuals with SCZ, however another study reported no sex differences^6^. Hypothalamic function is influenced by use of antipsychotic drugs; as an example, antipsychotic medication blocks the dopamine-2 (D_2_) receptor via the tuberoinfundibular dopamine pathway^34^. This pathway refers to a population of dopaminergic neurons in the ARC in the tuberal region of the hypothalamus. However, several studies reported no association between antipsychotic medication and hypothalamus volumes^5,6,7^.

In 2020, Billot et al^35^ released a method for the segmentation of the hypothalamus and its associated subnuclei. The results from the segmentation provide volumetric estimates of five subunits (and their corresponding nuclei) which are identifiable from visible landmarks based on standard resolution (∼1mm) T1-weighted MRI.^36,37.^ It is optimized for robustness across MRI scanner type and pulse sequence. Such automated methods enable the segmentation of hypothalamic volumes in large datasets while ensuring the consistency of the delineation of the hypothalamus and its subunits.

In the present study, we investigated diagnostic group differences in the segmented hypothalamus subunit volumes in a well-powered sample of 684 patients with SCZ or BPD and 684 age-and-sex-matched healthy controls. We further investigated differences and volumetric associations with age, sex, clinical symptoms, and psychotropic medication use. Given the role of the hypothalamus in the HPA axis, we hypothesized volumetric differences to occur in the PVN and SON, translating to lower volumes in the anterior-superior and anterior-superior subunit, in both SCZ and BPD. Given the role of dopaminergic neurons, as stated before, we hypothesized then volumetric differences in the ARC, influenced by the use of antipsychotic drugs. In the end, given previous reports of circadian rhythm disturbances^38^, we hypothesized that there would be volumetric differences in the SCN in individuals with SCZ or BPD compared to healthy controls. Due to the lack of previous studies on interactions with sex, and the paucity of studies on associations with medication use and clinical symptoms, we conducted these analyses on an exploratory basis.

## Materials and methods

### Participants

Participants were recruited as part of the ongoing Thematically Organized Psychosis (TOP) study (Oslo, Norway). Patients with a diagnosis of schizophrenia spectrum disorder (SCZ, n=344, mean age=33.3 (9.1), females=46.9%), including schizophrenia, schizophreniform and schizoaffective disorder, and individuals with a diagnosis of bipolar disorders (BPD, n=340, mean age=33 (11.3), females=57.9%), including bipolar I, bipolar II and bipolar disorder not otherwise specified, were referred from psychiatric outpatient clinics in the greater Oslo region. Healthy controls (CTR, n=684, mean age=31, (9.2) years, females=39.8%) were recruited from the same catchment area using the Norwegian national population register.

Diagnoses were established by trained clinical psychologists or physicians, using the Structured Clinical Interview for DSM-IV axis 1 disorders (SCID-IV). CTR were excluded if they had a history of substance abuse or dependency, or if they, or any first-degree relative, had experienced a severe psychiatric disorder. Both patients and CTR were excluded if they had a history of previous moderate or severe head injury, neurological disorders, or any medical conditions that could affect brain function or mental disability (defined as IQ<70). All participants gave written informed consent. The study was conducted in accordance with the Helsinki declaration and was approved by the Regional Committee for Medical Research Ethics and the Norwegian Data Protection Authority.

### Symptom and medication assessments

Clinical symptoms were assessed using the Positive and Negative Syndrome Scale (PANSS)^39^. We investigated associations with the symptom dimensions from the PANSS Wallwork consensus five-factor model: Positive (P1 + P3 + P5 + G9), disorganized (P2 + N5 + G11), negative (N1 + N2 + N3 + N4 + N6 + G7), excited (P4 + P7 + G8 + G14), and depressive (G2 + G3 + G6) symptom dimensions^40^.

We collected information on the use of antipsychotic, mood stabilizing (lithium and antiepileptics), and antidepressant medication. “Chlorpromazine equivalent (CPZ) doses were calculated using conversion values provided by Woods^41^.

We assessed current general intellectual level using a licensed translated version of the two-test version of the Wechsler Abbreviated Scale of Intelligence (WASI-II)^42^.

Age of onset was defined as the age at first psychotic episode for SCZ, and age at first affective episode, including manic, hypomanic, depressed, or mixed episodes for BPD.

### MRI acquisition

Participants underwent MRI scanning at the Core Facility MRI Neuroimaging, Oslo University Hospital, Ullevål, from 2005 to 2019, on three different scanners: 1.5T: 8-channel head coil; Siemens Magnetom Sonata (Siemens Medical Solutions, Erlangen, Germany) from 2005 to 2011; 3T General Electric (GE) Signa HDxt: 8-channel head coil from 2012 to 2014; 3T GE Discovery MR750: 32-channel head coil from 2015 to 2019. See **Supplementary 1** for an overview of the T1-weighted MRI pulse sequences used for each dataset.

### MRI processing

T1-weighted structural MRI images were first processed with FreeSurfer (v6.0.0; https://surfer.nmr.mgh.harvard.edu/). The Sequence Adaptive Multimodal SEGmentation (SAMSEG) tool, included in FreeSurfer v 7.2, was used to compute the segmentation-based Total Intracranial Volume (sbTIV).

Hypothalamic subunits were segmented using a Bayesian segmentation included in FreeSurfer v7.2^38^. This method estimates the volumes of five hypothalamic subunits in each hemisphere according to the grouping by Bocchetta et al.^36^ and Makris et al.^37^, in addition to the whole hypothalamus: 1) The anterior-inferior subunit (a-iHyp), which includes the suprachiasmatic and the supraoptic nucleus, 2) the anterior-superior subunit (a-sHyp), which includes the preoptic area and the paraventricular nucleus, 3) the posterior subunit (posHyp), comprised of the mammillary bodies (including medial and lateral mammillary nuclei), the lateral hypothalamus, and the tuberomammillary nucleus, 4) the inferior tubular subunit (infTub), which includes the arcuate nucleus, ventromedial nucleus, lateral tubular nucleus, and the tuberomammillary nucleus, and 5) the superior tubular subunit (supTub), which includes the dorsomedial nucleus, the paraventricular nucleus, and the lateral hypothalamus^38^. See **Table 2 and Figure 1**.

**Table 1.** Demographic and clinical data. Abbreviations: CTR: Healthy controls; BPD: Bipolar spectrum disorder; SCZ: Schizophrenia spectrum disorder; SD: standard deviation; IQ; Intelligence quotient estimate (from WASI). AOO: Age of onset; CPZ: Chlorpromazine equivalent dosage of antipsychotic medication. Comparison across groups with the Kruskal-Wallis test (KW) for non-normally distributed variables. The chi-square test (χ^2^) was used to compare categorical data. Age, IQ, education in years, and sex were compared between individuals with SCZ, BPD, and CTR. The PANSS Wallwork five-factors, age of onset, and CPZ-equivalent doses were compared between individuals with SCZ and BPD.

|  |  | **CTR**<br>**n= 684** | **BPD**<br>**n= 340** | **SCZ**<br>**n= 344** | **Statistical comparisons** |  |  | **Test** |
| --- | --- | --- | --- | --- | --- | --- | --- | --- |
| **Age** | *Mean, SD* | 33.3 (9.1) | 33 (11.3) | 31 (9.2) | CTR > SCZ<br>p = 1.599e-05 | CTR, BPD<br>p = 0.096 | BPD > SCZ<br>p = 0.039 | KW |
|  | *Range* | [18, 59.2] | [17.5, 65.8] | [18.5, 62.7] |  |  |  |  |
| **Female** | % | 46.9% | 57.9% | 39.8% | Diagnosis, Sex<br>p = 1.022e-05 | | | $\chi^2$ |
| **Education (y)** | *Mean, SD* | 14.4 (2.2) | 13.5 (2.3) | 12.7 (2.5) | CTR > SCZ<br>p < 2.2e-16 | CTR > BPD<br>p = 4.623e-09 | BPD > SCZ<br>p = 1.044e-06 | KW |
|  | *Range* | [9, 20] | [9, 20] | [4, 23] |  |  |  |  |
| **IQ** | *Mean, SD* | 113.8 (10.2) | 109.6 (12.1) | 102.9 (13.9) | CTR > SCZ<br>p < 2.2e-16 | CTR > BPD<br>p = 1.106e-07 | BPD > SCZ<br>p = 1.917e-09 | KW |
|  | *Range* | [75, 138] | [75, 137] | [70, 133] |  |  |  |  |
| **AOO** | *Mean, SD* |  | 20.8 (8.5) | 23.3 (7.2) | SCZ > BPD<br>p = 6.44e-11 |  |  | KW |
|  | *Range* |  | [7, 55] | [7, 51] |  |  |  |  |
| **Wallwork factors** |  |  |  |  |  |  |  |  |
| **Positive** | *Mean, SD* |  | 5.6 (2.5) | 9.5 (4.1) | SCZ > BPD<br>p < 2.2e-16 |  |  | KW |
|  | *Range* |  | [4, 22] | [4, 23] |  |  |  |  |
| **Negative** | *Mean, SD* |  | 8.6 (3.3) | 13.3 (5.9) | SCZ > BPD<br>p < 2.2e-16 |  |  | KW |
|  | *Range* |  | [6, 22] | [6, 34] |  |  |  |  |
| **Excited** | *Mean, SD* |  | 5.1 (1.6) | 5.4 (2.03) | SCZ, BPD<br>p = 0.093 |  |  | KW |
|  | *Range* |  | [4, 16] | [4, 19] |  |  |  |  |
| **Depressed** | *Mean, SD* |  | 7.7 (2.9) | 8.1 (3.1) | SCZ, BPD<br>p = 0.077 |  |  | KW |
|  | *Range* |  | [3, 18] | [3, 16] |  |  |  |  |
| **Disorganized/concrete** | *Mean, SD* |  | 4.0 (1.5) | 5.4 (2.6) | SCZ > BPD<br>p < 2.2e-16 |  |  | KW |
|  | *Range* |  | [3, 12] | [3, 19] |  |  |  |  |
| **CPZ** | *Mean, SD* |  | 216.8 (187.86) | 359.62 (260.38) | SCZ > BPD<br>p < 2.2e-16 |  |  | KW |
|  | *Range* |  | [17.61, 1288.95] | [36.01, 2485.02] |  |  |  |  |
| **Lithium Yes** | % |  | 17% (3 NA's) | 3.8% |  |  |  |  |
| **Antidepressants Yes** | % |  | 35% (3 NA's) | 29.4% |  |  |  |  |
| **Antipsychotics Yes** | % |  | 49% (3 NA's) | 89.1% |  |  |  |  |
| **Antiepileptics Yes** | % |  | 37% (3 NA's) | 13.8% |  |  |  |  |

**Table 2.** Grouping of the hypothalamic nuclei into subunits. ^35,36,37^.

| Subunit | Nuclei included |
| --- | --- |
| Anterior-superior (a-sHyp) | preoptic area; paraventricular nucleus (PVN) |
| Anterior-inferior (a-iHyp) | suprachiasmatic nucleus; supraoptic nucleus (SON) |
| Superior tubular (supTub) | dorsomedial nucleus; PVN; lateral hypothalamus |
| Inferior tubular (infTub) | infundibular (or arcuate) nucleus; ventromedial nucleus; SON; lateral tubular nucleus; tuberomamillary nucleus (TMN) |
| Posterior (posHyp) | mamillary body (including medial and lateral mamillary nuclei); lateral hypothalamus; TMN |

**Figure 1.**
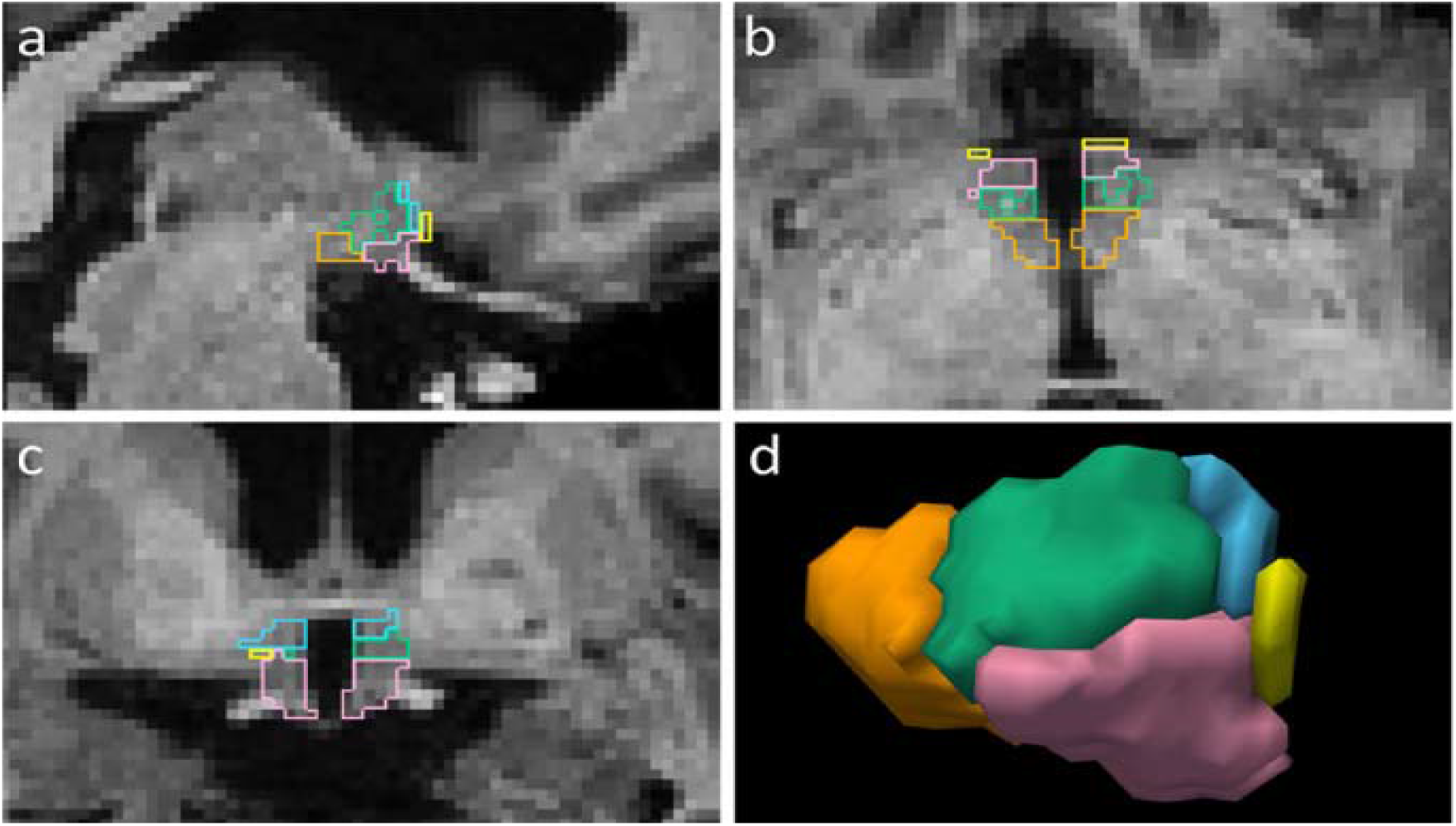
Example of manually segmented hypothalamus in (a) sagittal, (b) axial and (c) coronal views. (d) 3D rendering of the right hypothalamus. Subunits are depicted in different colours: a-sHyp in blue, a-iHyp in yellow, supTub in green, infTub in pink, and posHyp in orange. [Image from Billot et al, 2020^35^].

ComBat^43,44^ was used to harmonize whole hypothalamus and hypothalamic subunit volumes, as well as sbTIV, across the three different MRI platforms. We entered sex, age, and diagnosis as biological variables of interest.

### Statistical analyses

Statistical analyses were performed with R version 4.2.1 (R Core Team 2020). To correct for multiple testing, we used the Benjamini-Hochberg procedure^45^. Unless otherwise specified, adjusted p-values are reported. Cohen’s d effect sizes and their 95% confidence intervals based on t-values were computed using the *effectsize* package^46^. CTR were age- and sex-matched to the combined patient group using nearest neighbor matching with the *matchit* function implemented in R^47^.

#### 1. Main analyses

In the main analysis, we compared individuals with SCZ and BPD to CTR by fitting regression models with total hypothalamus and hypothalamic subunit volumes, for each hemisphere, as dependent variables. These models were covaried for age, age^2^, sex, and sbTIV, with diagnostic group as the variable of interest.

#### 2. Effects of age and sex

To investigate putative age-by-diagnosis and sex-by-diagnosis interaction effects, we fitted separate regression models as specified above, but additionally including age-by-diagnosis and sex-by-diagnosis interaction terms, respectively. Where sex-by-diagnosis was significant, we followed up with sex-stratified analyses, where the regression models described above were fitted for male and female participants separately, thus comparing female patients with female CTR and male patients with male CTR.

#### 3. Associations with clinical symptoms and age of onset

To test for associations with clinical symptoms and age of onset, we fitted regression models for SCZ and BPD separated. These models were covaried for age, age^2^, sex, and sbTIV. As variables of interest, we included age of onset and the five Wallwork factors, i.e., the positive, disorganized, negative, excited, and depressive symptom dimensions.

#### 4. Associations with psychotropic medication

We tested for associations with psychotropic medication by fitting regression models with the same covariates as above for each diagnostic group separately. Here, we included as variables of interest antipsychotic medication use (yes/no) as well as CPZ-equivalent dosage among users, antidepressant use (yes/no), antiepileptic use (yes/no). For individuals with BPD, we additionally tested for associations with lithium use (yes/no).

#### 5. Post hoc analyses

To investigate if and how ComBat harmonization affected the results, we fitted the same regression models as in the main analyses with the non-ComBat harmonized volumes. This was done for the subunits where significant differences were found in the main analysis. Further, we fitted the main models with ComBat harmonized volumes, for the subunits that were significantly different in the main analysis, with scanner as an additional covariate.

## Results

### 1. Main analyses

For individuals with SCZ, we found larger left infTub (p=0.034; d=0.16) volume, as well as smaller volumes of the right a-iHyp (p=0.0003; d=-0.24), a-sHyp (p=0.034; d=-0.16), and posHyp (p<0.05; d=-0.15), compared to CTR. No group differences between individuals with BPD and CTR remained significant after correction for multiple testing (see **Figure 2 & 3** and supplementary **Figure S1 and Table S1**).

**Figure 2.**
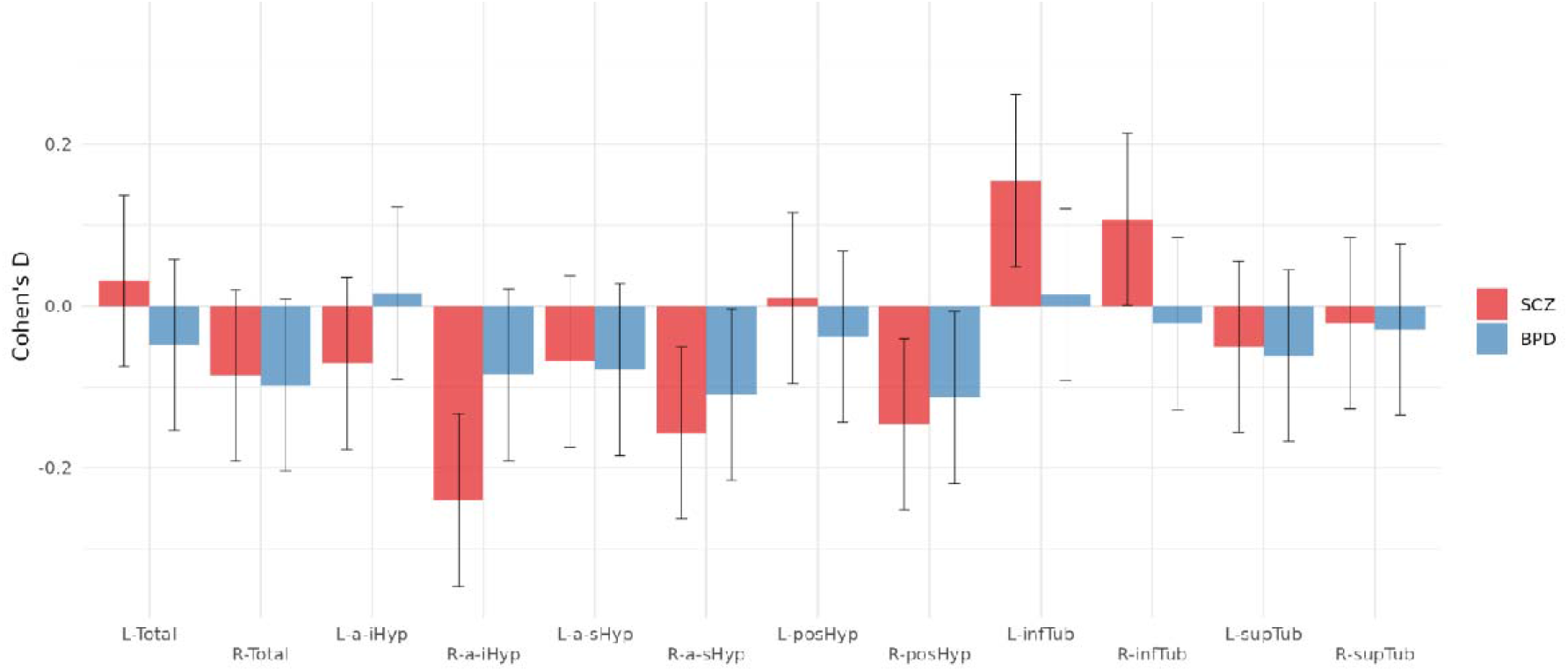
Cohen’s d effect sizes for schizophrenia spectrum disorders (SCZ) and bipolar disorders (BPD) compared to healthy controls (CTR) for each hypothalamic subunit. Error bars show the 95% confidence intervals of the estimated effect sizes. Figures were made with the *ggplot* package^69^.

**Figure 3.**
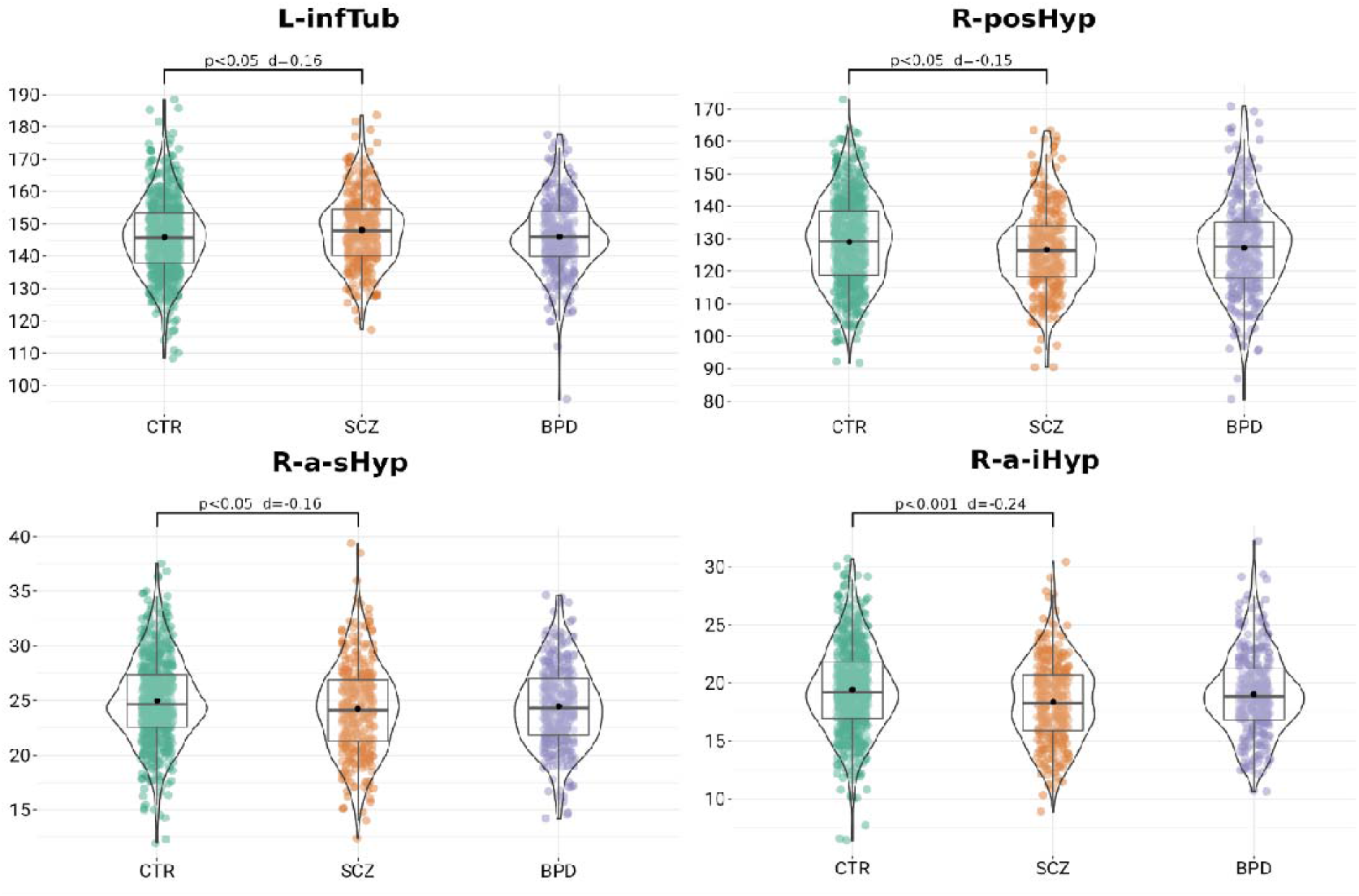
Violin plots of residualized volumes centered around the grand mean for each diagnostic group and region that was significantly different in the group analysis. Abbreviations: L-infTub= Left inferior tubular subunit; R-posHyp=right posterior subunit; R-a-sHyp=right anterior superior subunit; R-a-iHyp=right anterior inferior subunit.

### 2. Effects of age and sex

None of the age-by-diagnostic group interactions were statistically significant after correction for multiple testing. No sex-by-SCZ/CTR interaction was significant. Significant sex-by-BPD/CTR interactions were observed for the whole hypothalamus in both the left (p=0.047; d=-0.16) and right (p=0.047; d=-0.16) hemispheres.

In a sex-stratified analysis on the whole right hypothalamus, males with BPD had significantly smaller volumes compared to CTR males (p=0.038; d=-0.22), whereas females with BPD did not differ from CTR females. Also, males with BPD had a smaller right posHyp (p=0.044; d=-0.21) compared to CTR. In a sex-stratified analysis on the whole left hypothalamus and left hypothalamic nuclei males and females with BPD did not differ from CTR.

### 3. Associations with clinical symptoms and age of onset

There were no statistically significant associations between hypothalamic volumes and any of the five Wallwork factors or with the age of onset.

### 4. Associations with psychotropic medication

We found significantly larger volume of the left tubInf (p=0.039; d=0.33) in individuals with BPD currently on antipsychotics compared to those not currently on antipsychotics. No other associations were found between hypothalamic volumes and psychotropic medication.

### 5. Post hoc analyses

The significant differences between SCZ and CTR from the main analysis, i.e., the left infTub and the right a-iHyp, a-sHyp, and posHyp, remained significant both without ComBat harmonization and after adding scanner as an additional covariate.

## Discussion

Individuals with SCZ showed a complex pattern of regional alterations within the hypothalamus: while the left inferior tubular (infTub) subunit was larger when compared with CTR, the right anterior-inferior (a-iHyp), right anterior-superior (a-sHyp), and right posterior subunits (p-Hyp) were smaller. It is worth noting that this pattern was found although the total hypothalamic volume did not differ compared with the CTR, and that it was found in SCZ but not in BPD.

Previous postmortem studies^48,49^ did not find volumetric differences in these regions; however, these studies had small samples (approx. n=10 in each group) and may have been underpowered to detect subtle morphometric effects. To our knowledge, our study is the first to perform a large-scale examination of hypothalamic subregions *in vivo* in this patient group.

Smaller a-iHyp subunit volumes in SCZ indicate alterations in the *suprachiasmatic nucleus* (SCN) and *supra-optic nucleus* (SON). The SCN controls endogenous circadian rhythms^50^ and there is emerging evidence that dysfunction of the SCN and the associated sleep disturbances may play an important role in clinical manifestations of SCZ ^51^. There is also evidence that dopamine could influence the circadian clock, through entrainment of the master clock in the SCN^14^. However, BPD are also associated with circadian rhythm disruption, and we did not find smaller or altered a-iHyp volume among individuals with BPD.

The SON, together with *paraventricular nucleus* (PVN), produce nonapeptide oxytocin and arginine–vasopressin. Arginine-vasopressin is also expressed in SCN neurons^52^. Arginine-vasopressin and oxytocin are known to be involved in the modulation of social interaction, and and lower blood plasma levels of these hormones were reported in a study of male patients with SCZ^53^. Reduced level of arginine-vasopressin has been observed in SCZ, although the findings are not consistent, and lower expression of mRNA encoding vasopressin has been reported in the PVN, but not the SON, however, the pathophysiological relevance remains to be clarified^54^.

We further observed a smaller a-sHyp subunit volume in the right hemisphere in individuals with SCZ compared with CTR. Contrary, Goldstein and colleagues^6^ observed larger anterior hypothalamus volumes, including the PVN, in SCZ. Since PVN has a high density of corticotropin-releasing hormone (CRH), implicated in the stress response, this finding can be related to an alteration of the HPA-axis. CRH neurons in the PVN colocalize with sex-hormone receptors, such as estrogen receptors (ER-alpha and ER-beta), suggesting a role for ERs in the PVN release of CRH^55^. Several studies have also reported higher rates of baseline cortisol activity in SCZ and hyperresponsivity to stress^56,57,58-6^.

In addition, for the right p-Hyp subunit, which includes the *mammillary bodies*, we observed smaller volumes in individuals with SCZ compared to CTR. Previous findings showed conflicting results, but in an early study on postmortem brain, mammillary bodies volume was also smaller in SCZ relative to CTR ^59^. Mammillary bodies may be important neuronal-relay centers in the temporolimbic system, implicated in memory function^60^.

Neuropsychological studies have consistently shown global cognitive dysfunction as well as specific memory impairments in SCZ^61^. Episodic memory deficits, in particular during retrieval conditions with high cognitive demand, have been demonstrated in tasks focusing on cognitive control, relational encoding and intentional forgetting^62^.

In individuals with SCZ, we also observed larger left infTub volumes compared to CTR. The *Arcuate nucleus* (ARC) is part of the infTub subunits. Dopamine and growth hormone-releasing hormone are synthesized in the ARC, and prolactin, released from the anterior pituitary, is controlled by dopamine^27^. While often associated with antipsychotic medication use, hyperprolactinemia can also be stress-induced in individuals with SCZ and, since elevated prolactin levels can increase dopamine release through a feedback mechanism, this could contribute to explaining how stress can trigger the outbreak of psychosis^63^.

### Sex-stratified results

Although we found no significant interactions in the sex-by-SCZ/CTR analysis, we found a significant sex interaction in BPD for the total right and left hypothalamus. The sex-stratified analysis in BPD revealed smaller right hypothalamus volumes and right postHyp volumes in males with BPD compared to male CTR. No differences were found for females with BPD relative to same-sex CTR. Previous work in healthy populations suggest sex differences in hypothalamus volumes. For instance, Makris and colleagues found larger total hypothalamus volumes in males compared to females, after correction for head size^37^. In another study, male CTR had significantly larger mammillary bodies compared to females^64^. Plotting male vs female estimates for the hypothalamic nuclei, we observed a consistent difference in both the right and the left hemisphere of the hypothalamus and of its subnuclei (except both right and left a-iHyp subunit) between CTR males and CTR females (**Supplementary Figure 2**). In a previous study, it was demonstrated that in female CTR sex differences in anterior hypothalamus, including PVN and VMN, under stress, were dependent on gonadal hormone changes over the menstrual cycle ^65^. SON, PVN and ARC subunits are related to the HPA-axis, and they are involved in estradiol activity, which differs between males and females^66^. Further investigation of hormonal changes related to HPA-axis in association with specific gene polymorphism variation or different gene expression in line with the sex-stratified analysis, considering both CTR and patients, is needed.

### Associations with antipsychotics

In patients with BPD, antipsychotic treatment was associated with larger volumes in the left tubInf subunit. Within this region, the ARC contains dopaminergic neurons that are functionally involved in the homeostatic regulation of prolactin through the tubero-infundibular pathway. Thus, one possibility is that volumetric increase may occur as a compensatory response to dopaminergic antagonism, in line with previous observations of basal ganglia enlargement following antipsychotic treatment^67^. However, the VMN is also found in this region and is involved in metabolic pathways, specifically related to satiety and caloric intake. Thus, larger infTub volumes in antipsychotic-treated patients is a finding that may be relevant to pathways underlying two well-known side effects of antipsychotics: prolactin increases and weight gain. There was, however, no association with antipsychotic medication use in SCZ. As mentioned above, hyperprolactinemia has been demonstrated also in antipsychotic-naïve individuals with first episode SCZ^55^, suggesting that this may not always be a side effect. Still, it is worth noting that in the present study, previous medication use was not controlled for and that only a small proportion of individuals with SCZ (10.9%) were not treated with antipsychotic drugs at the time of examination. Further investigation of the relationship between infTub volumes and medication use, which takes the biological pathways discussed above into account may be warranted.

#### Strength and limitations

A major strength of the present study is the large and well-characterized cohort of patients with SCZ and BPD. We used a recently developed method for automatic segmentation of the hypothalamus which has shown high consistency even compared to human intra-rater precision. However, some limitations need to be considered. In our analyses, we pooled data from three different scanners. Although the hypothalamic subunit segmentation method has been optimized for robustness across scan platform and pulse sequence and we harmonized between scan platforms using ComBat, residual effects of scanner may have remained. Furthermore, the image resolution was low (approximately 1mm3 voxel resolution) relative to the size of the segmented subunits, which may have affected the accuracy of the volumetric estimates and did not allow us to examine volumetric differences at the level of individual nuclei.

## Conclusion

Our findings indicate that SCZ is associated with regionally specific structural alterations of the hypothalamus, which are present in the absence of altered whole hypothalamus volume. BPD is not associated with the same pattern of structural alterations, although in male patients we found significantly altered volumes.

## Data Availability

The data was collected by the Norwegian Research Centre for Mental Disorders (NORMENT) at the Oslo University Hospital (OUS). The data is subject to restrictions and is not publicly available but may be made available given reasonable request. Data can only be made available following permission from OUS, and insofar requests are in line with the relevant ethical agreements and the consent of the participants.

## Acknowledgements

We thank the study participants and the clinicians at the Norwegian Research Centre for Mental Disorders (NORMENT) who were involved in recruitment and assessments. Data services were provided by the “Tjeneste for Sensitive Data” (TSD) facilities, operated and developed by the TSD service group at the University of Oslo (USIT).

