## Supplementary 1; Supplementary figure 1, Supplementary table 1, Supplementary figure 2 for "Hypothalamic subunit volumes in schizophrenia and bipolar spectrum disorders"

**Supplementary 1. MRI pulse sequence acquisition parameters.**

Participants examined from 2005 to 2011 (n=666) were scanned using a 1.5T Siemens Magnetom Sonata (Siemens Medical Solutions, Erlangen, Germany) platform equipped with an eight-channel head coil. Sagittal T1-weighted magnetization prepared rapid gradient echo (MPRAGE) volumes were acquired with the following parameters: Echo time (TE) = 3.93 ms. Repetition time (TR) = 2730 ms. Inversion time (TI) = 1000 ms. Flip angle (FA) = 7°. Field of view (FOV) = 240mm, acquisition matrix: 192 x 256. Resolution (voxel size) = 1.33 x 0.94 x 1 mm^3^. To increase the signal-to-noise ratio (SNR), two volumes were acquired per subject and subsequently averaged, after rigid-body registration.

Participants examined during the time period 2012-2014 (n=383) were scanned using a 3T General Electric (GE) Signa HDxt platform equipped with an eight channel head coil. T1-weighted 3D Fast Spoiled Gradient Echo (FSPGR) volumes were acquired using the following parameters: TE= MinFull. TR=7.8ms. TI = 450 ms. FA = 12°. FOV = 256mm, matrix: 256 x 192. Resolution (voxel size) = 1 x 1 x 1.2 mm^3^.

Participants examined during the time period 2015-2019 (n=433) were scanned using a GE Discovery MR750 platform equipped with a 32 channel head coil. Inversion recovery-prepared 3D gradient recalled echo (3D BRAVO) volumes were acquired with the following parameters: TE = 3.18 ms. TR = 8.16 ms. TI = 400 ms. FA = 12°. FOV = 256mm, matrix: 188 x 256. Resolution (voxel size) = 1 x 1 x 1 mm^3^.

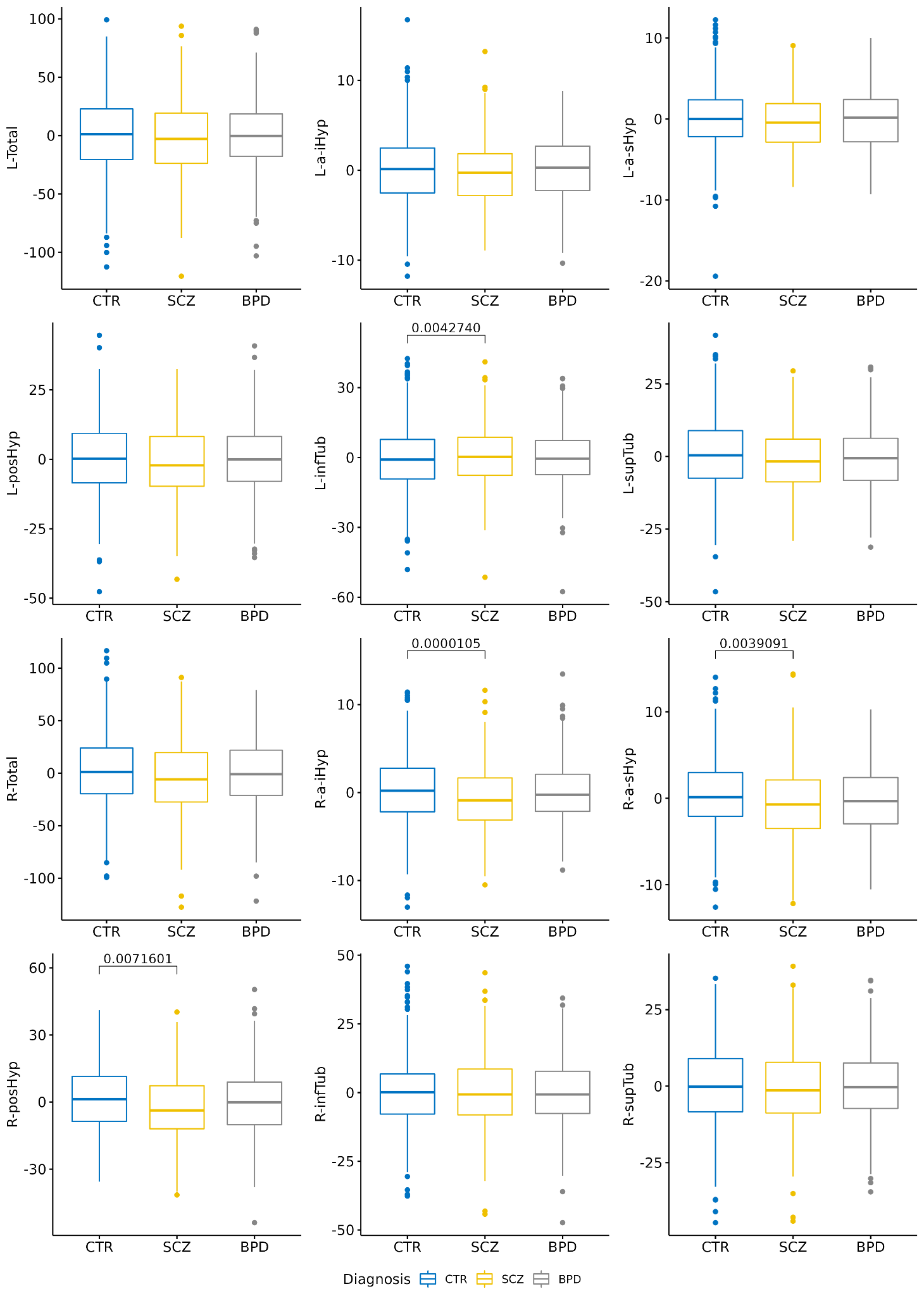

**Supplementary Figure 1.** Boxplots depicting residualized hypothalamic volumes for patients with SCZ, BPD, and healthy controls. Significant differences are labeled with uncorrected p-values

| **L-Total** | **Estimate** | **Std. Error** | **t value** | **Pr(>\|t\|)** |
| --- | --- | --- | --- | --- |
| (Intercept) | 167.453 | 12.568 | 13.324 | <0.0001 |
| SexMale | 18.285 | 1.890 | 9.674 | <0.0001 |
| Age | 0.886 | 0.461 | 1.923 | 0.055 |
| I(Age^2) | -0.015 | 0.006 | -2.376 | 0.018 |
| sbTIV_ComBat | 0.0001 | <0.0001 | 24.341 | <0.0001 |
| DiagnosisSCZ | 0.996 | 1.749 | 0.570 | 0.569 |
| DiagnosisBPD | -1.555 | 1.760 | -0.884 | 0.377 |

| **L-a-iHyp** | **Estimate** | **Std. Error** | **t value** | **Pr(>\|t\|)** |
| --- | --- | --- | --- | --- |
| (Intercept) | 10.952 | 1.728 | 6.340 | <0.0001 |
| SexMale | 0.141 | 0.260 | 0.541 | 0.588 |
| Age | 0.075 | 0.063 | 1.192 | 0.233 |
| I(Age^2) | -0.0008 | 0.0009 | -0.959 | 0.338 |
| sbTIV_ComBat | <0.0001 | <0.0001 | 4.490 | <0.0001 |
| DiagnosisSCZ | -0.312 | 0.240 | -1.297 | 0.195 |
| DiagnosisBPD | 0.071 | 0.242 | 0.295 | 0.768 |

| **L-a-sHyp** | **Estimate** | **Std. Error** | **t value** | **Pr(>\|t\|)** |
| --- | --- | --- | --- | --- |
| (Intercept) | 12.951 | 1.735 | 7.464 | <0.0001 |
| SexMale | 1.4624 | 0.261 | 5.604 | <0.0001 |
| Age | 0.067 | 0.064 | 1.057 | 0.291 |
| I(Age^2) | -0.001 | 0.0009 | -1.158 | 0.247 |
| sbTIV_ComBat | <0.0001 | <0.0001 | 7.774 | <0.0001 |
| DiagnosisSCZ | -0.305 | 0.241 | -1.263 | 0.207 |
| DiagnosisBPD | -0.351 | 0.243 | -1.447 | 0.148 |

| **L-posHyp** | **Estimate** | **Std. Error** | **t value** | **Pr(>\|t\|)** |
| --- | --- | --- | --- | --- |
| (Intercept) | 31.883 | 5.636 | 5.657 | <0.0001 |
| SexMale | 3.137 | 0.848 | 3.701 | 0.0002 |
| Age | 0.598 | 0.207 | 2.897 | 0.004 |
| I(Age^2) | -0.009 | 0.003 | -3.073 | 0.0022 |
| sbTIV_ComBat | <0.0001 | <0.0001 | 18.606 | <0.0001 |
| DiagnosisSCZ | 0.144 | 0.784 | 0.183 | 0.855 |
| DiagnosisBPD | -0.554 | 0.789 | -0.702 | 0.483 |

| **L-infTub** | **Estimate** | **Std. Error** | **t value** | **Pr(>\|t\|)** |
| --- | --- | --- | --- | --- |
| (Intercept) | 58.411 | 5.548 | 10.528 | <0.0001 |
| SexMale | 6.434 | 0.834 | 7.711 | <0.0001 |
| Age | -0.064 | 0.203 | -0.313 | 0.755 |
| I(Age^2) | 0.0008 | 0.003 | 0.292 | 0.771 |
| sbTIV_ComBat | <0.0001 | <0.0001 | 18.517 | <0.0001 |
| DiagnosisSCZ | 2.209 | 0.772 | 2.862 | 0.004 |
| DiagnosisBPD | 0.202 | 0.777 | 0.260 | 0.795 |

| **L-supTub** | **Estimate** | **Std. Error** | **t value** | **Pr(>\|t\|)** |
| --- | --- | --- | --- | --- |
| (Intercept) | 51.442 | 5.312 | 9.683 | <0.0001 |
| SexMale | 6.823 | 0.799 | 8.539 | <0.0001 |
| Age | 0.212 | 0.195 | 1.089 | 0.276 |
| I(Age^2) | -0.005 | 0.003 | -1.988 | 0.047 |
| sbTIV_ComBat | <0.0001 | <0.0001 | 14.926 | <0.0001 |
| DiagnosisSCZ | -0.680 | 0.739 | -0.919 | 0.358 |
| DiagnosisBPD | -0.835 | 0.744 | -1.122 | 0.262 |

| **R-Total** | **Estimate** | **Std. Error** | **t value** | **Pr(>\|t\|)** |
| --- | --- | --- | --- | --- |
| (Intercept) | 164.883 | 13.940 | 11.828 | <0.0001 |
| SexMale | 16.223 | 2.097 | 7.738 | <0.0001 |
| Age | 1.030 | 0.511 | 2.017 | 0.044 |
| I(Age^2) | -0.020 | 0.007 | -2.928 | 0.003 |
| sbTIV_ComBat | 0.0007 | <0.0001 | 21.486 | <0.0001 |
| DiagnosisSCZ | -3.073 | 1.939 | -1.585 | 0.113 |
| DiagnosisBPD | -3.511 | 1.952 | -1.799 | 0.072 |

| **R-a-iHyp** | **Estimate** | **Std. Error** | **t value** | **Pr(>\|t\|)** |
| --- | --- | --- | --- | --- |
| (Intercept) | 13.164 | 1.758 | 7.490 | <0.0001 |
| SexMale | 0.877 | 0.264 | 3.318 | 0.0009 |
| Age | 0.102 | 0.064 | 1.587 | 0.113 |
| I(Age^2) | -0.001 | 0.0008 | -1.712 | 0.087 |
| sbTIV_ComBat | <0.0001 | <0.0001 | 2.855 | 0.004 |
| DiagnosisSCZ | -1.081 | 0.244 | -4.423 | <0.0001 |
| DiagnosisBPD | -0.388 | 0.246 | -1.576 | 0.115 |

| **R-a-sHyp** | **Estimate** | **Std. Error** | **t value** | **Pr(>\|t\|)** |
| --- | --- | --- | --- | --- |
| (Intercept) | 14.990 | 1.900 | 7.889 | <0.0001 |
| SexMale | 1.344 | 0.286 | 4.703 | <0.0001 |
| Age | 0.050 | 0.067 | 0.718 | 0.473 |
| I(Age^2) | -0.001 | 0.0009 | -1.440 | 0.150 |
| sbTIV_ComBat | <0.0001 | <0.0001 | 5.860 | <0.0001 |
| DiagnosisSCZ | -0.764 | 0.264 | -2.890 | 0.004 |
| DiagnosisBPD | -0.534 | 0.266 | -2.008 | 0.045 |

| **R-posHyp** | **Estimate** | **Std. Error** | **t value** | **Pr(>\|t\|)** |
| --- | --- | --- | --- | --- |
| (Intercept) | 40.171 | 6.519 | 6.163 | <0.0001 |
| SexMale | 2.417 | 0.980 | 2.465 | 0.014 |
| Age | 0.709 | 0.239 | 2.967 | 0.003 |
| I(Age^2) | -0.012 | 0.003 | -3.662 | 0.0002 |
| sbTIV_ComBat | <0.0001 | <0.0001 | 14.458 | <0.0001 |
| DiagnosisSCZ | -2.443 | 0.907 | -2.693 | 0.007 |
| DiagnosisBPD | -1.903 | 0.913 | -2.085 | 0.037 |

| **R-infTub** | **Estimate** | **Std. Error** | **t value** | **Pr(>\|t\|)** |
| --- | --- | --- | --- | --- |
| (Intercept) | 48.666 | 5.339 | 9.114 | <0.0001 |
| SexMale | 5.670 | 0.803 | 7.060 | <0.0001 |
| Age | -0.086 | 0.196 | -0.439 | 0.660 |
| I(Age^2) | 0.0004 | 0.003 | 0.158 | 0.875 |
| sbTIV_ComBat | <0.0001 | <0.0001 | 19.388 | <0.0001 |
| DiagnosisSCZ | 1.470 | 0.743 | 1.978 | 0.048 |
| DiagnosisBPD | -0.296 | 0.748 | -0.396 | 0.692 |

| **R-supTub** | **Estimate** | **Std. Error** | **t value** | **Pr(>\|t\|)** |
| --- | --- | --- | --- | --- |
| (Intercept) | 46.038 | 5.602 | 8.219 | <0.0001 |
| SexMale | 5.668 | 0.842 | 6.728 | <0.0001 |
| Age | 0.284 | 0.205 | 1.385 | 0.166 |
| I(Age^2) | -0.006 | 0.003 | -2.288 | 0.0222 |
| sbTIV_ComBat | <0.0001 | <0.0001 | 15.602 | <0.0001 |
| DiagnosisSCZ | -0.304 | 0.779 | -0.390 | 0.697 |
| DiagnosisBPD | -0.417 | 0.784 | -0.532 | 0.595 |

**Supplementary Table 1.** Output from the main analysis. Comparison between patients with SCZ (schizophrenia) and BPD (bipolar disorder) with controls, by fitting regression models with total hypothalamus and hypothalamic subunit volumes, for both hemispheres, as dependent variables. These models were covaried for age, age^2^, sex, and sbTIV, with diagnostic group as the variable of interest.

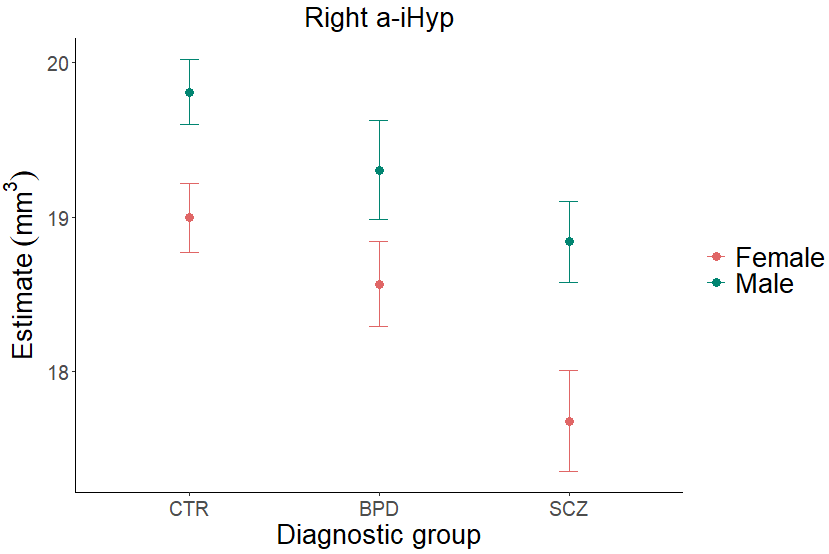

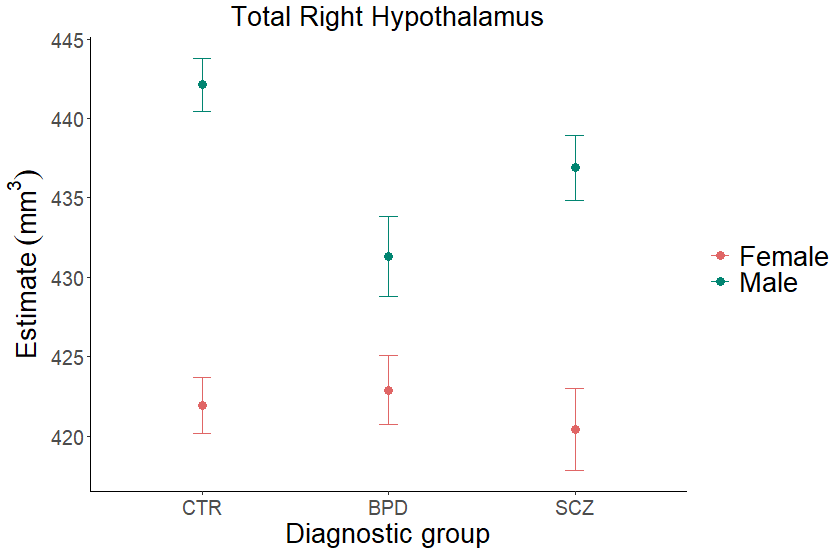

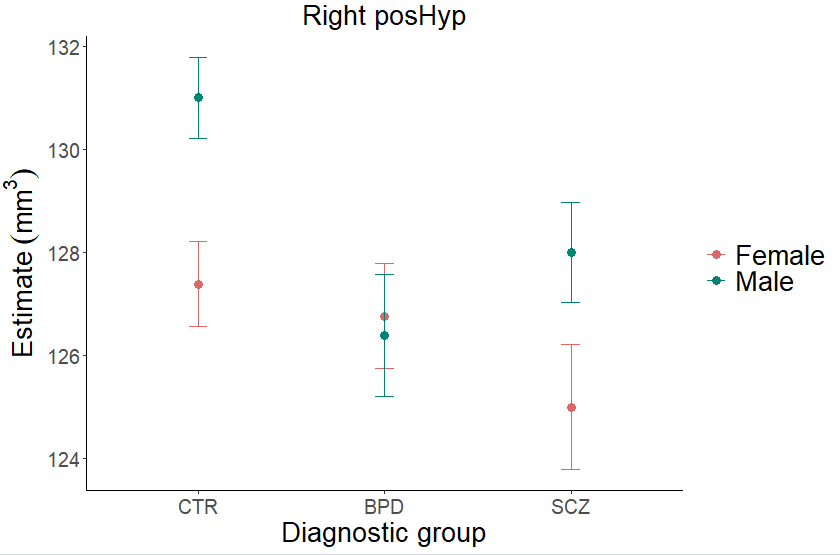

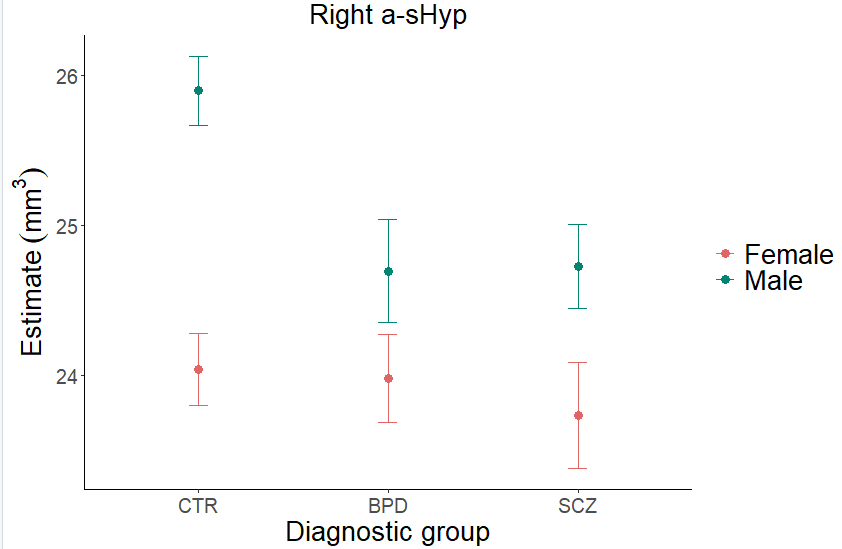

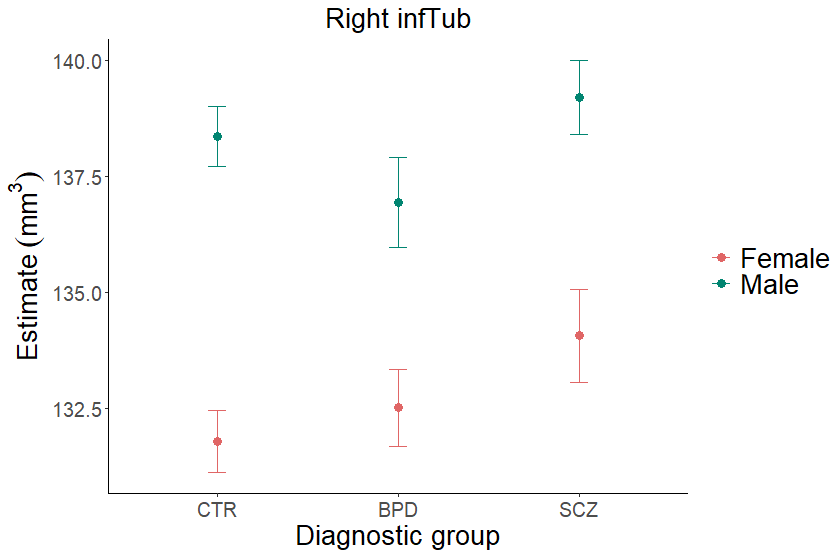

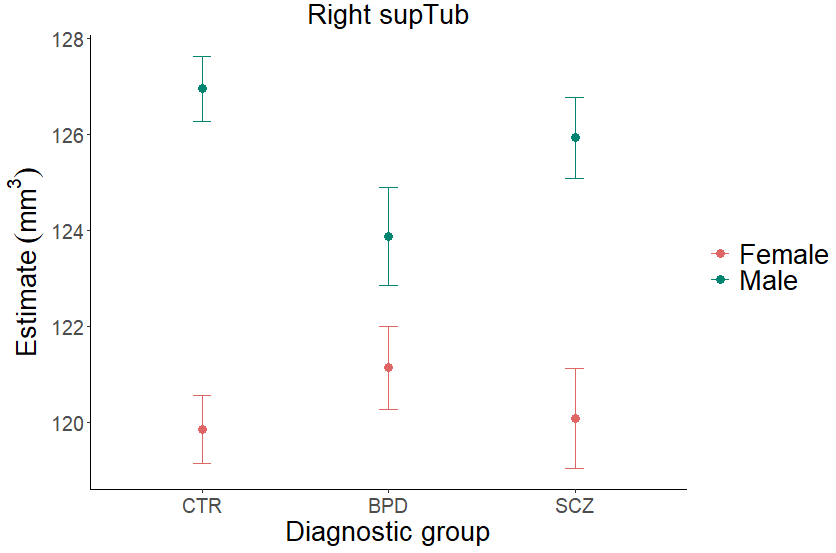

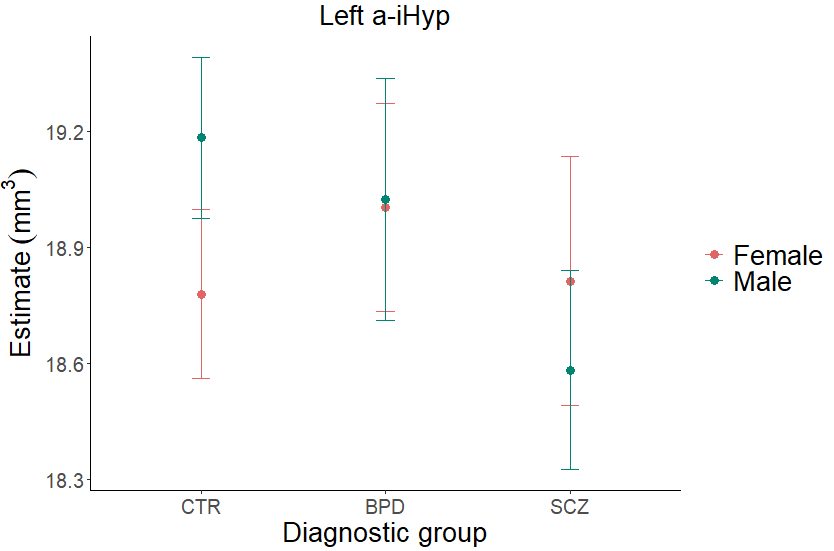

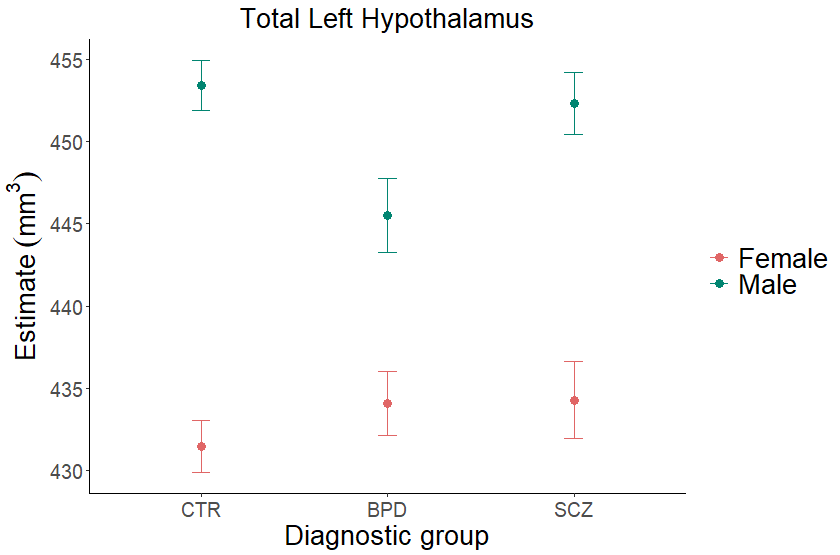

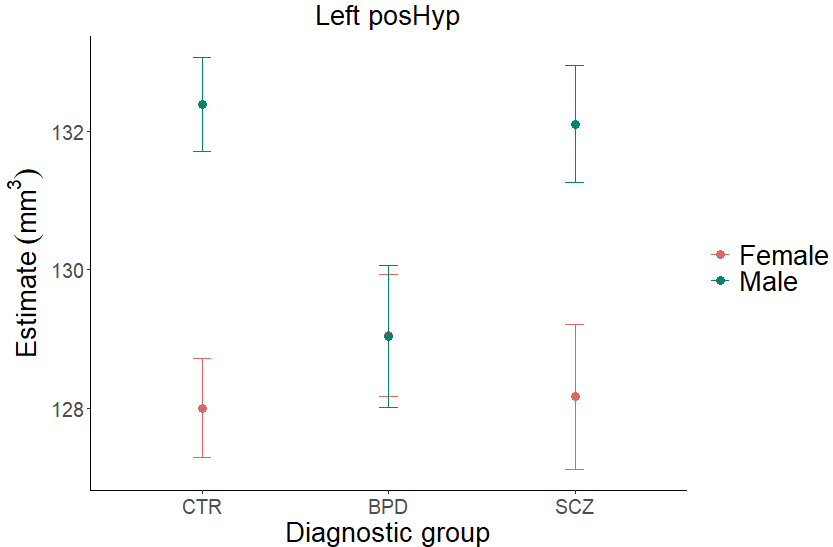

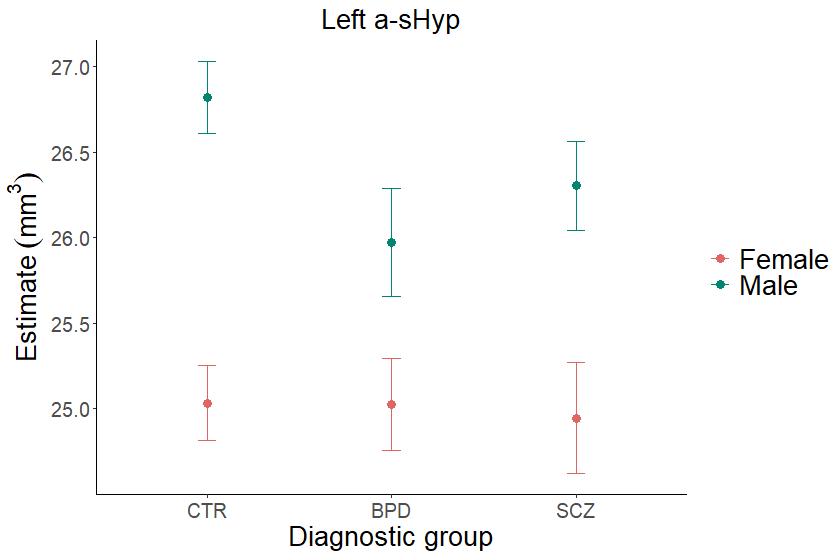

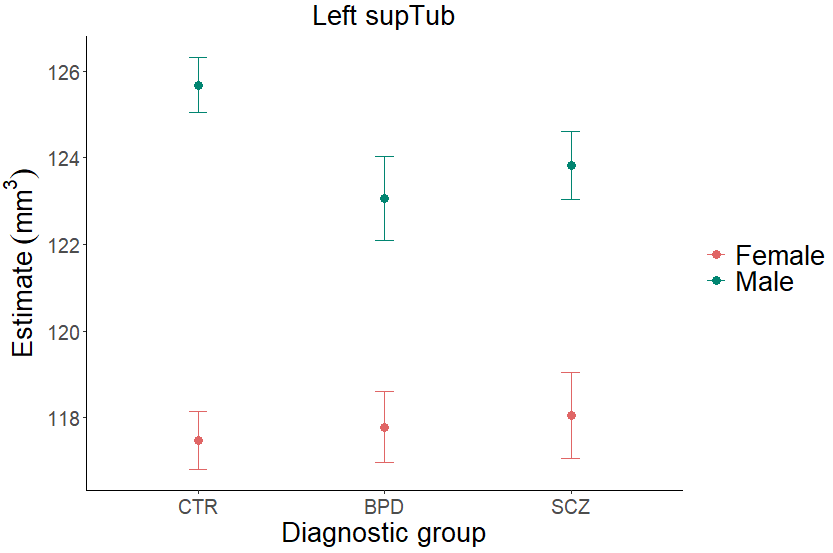

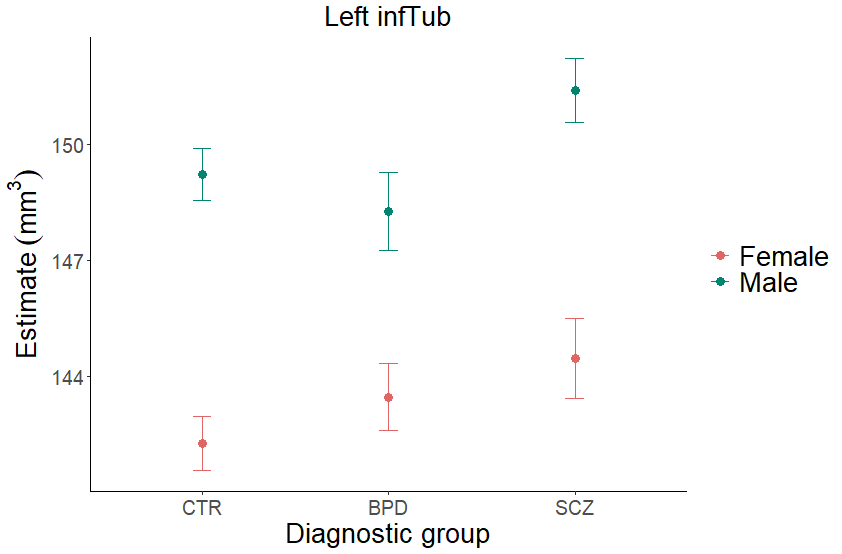

**Supplementary Figure 2.** Estimates of hypothalamic volumes stratified by diagnostic subgroup. Estimates are displayed with upper and lower CIs, adjusted for for age, age^2^, sex, and sbTIV. CTR = healthy controls BPD = bipolar disorder, SCZ = schizophrenia.
